# Accelerated long-term forgetting as an objective marker of subjective memory impairment in multiple sclerosis

**DOI:** 10.64898/2026.04.21.26351393

**Authors:** Christina Jansen, Johannes Stalter, Sigrid Reuter, Karsten Witt

## Abstract

**Background:** Accelerated long-term forgetting (ALF), defined as an increased rate of memory loss over extended intervals, has so far been detected in a pilot study of patients with mild multiple sclerosis (MS). This study aimed to (I) confirm the presence of ALF in a larger, heterogeneous MS sample, (II) explore associations with patient-reported outcomes, and (III) assess the diagnostic performance of ALF tests for subjective memory impairment.

**Methods:** This study compared 62 MS patients and 65 age-, sex-, and education-matched healthy controls using standardized memory tests (RAVLT, WMS-IV Logical Memory subtest). Recall was assessed immediately, after 30 minutes, and after 7 days. Seven-day/30-minute recall ratios (Q_RAVLT_, Q_WMS_) served as primary outcomes. Self-report measures included memory complaints, fatigue, depression, and sleep disturbances. Linear regression and Receiver operating characteristic (ROC) analyses assessed predictors and diagnostic accuracy.

**Results:** ALF was observed in multiple sclerosis since Q_RAVLT_ was lower in patients than in controls (0.64 [95% CI 0.59–0.69] vs. 0.78 [0.73–0.82], p < 0.001), as was Q_WMS_ (0.79 [95% CI 0.74–0.84] vs. 0.95 [0.90–1.00], p < 0.001), despite comparable initial learning. Greater fatigue, higher memory complaints, longer disease duration, older age, and greater disability were associated with lower ALF scores. The combined ALF score moderately discriminated subjective memory impairment (AUC 0.74; sensitivity 0.73; specificity 0.73).

**Conclusion:** MS patients showed ALF despite normal initial learning, indicating a specific memory deficit undetected by standard tests. Long-delay recall using RAVLT and WMS-IV Logical Memory subtest may improve cognitive impairment detection in MS.

## 1 Introduction

Cognitive impairments, particularly memory deficits, are common in multiple sclerosis (MS), a chronic autoimmune-mediated neurological disease (Benedict et al., 2020). These deficits can substantially affect patients’ quality of life, yet in some cases they can be difficult to detect in routine clinical assessments (Amato et al., 2006). A specific type of memory impairment, accelerated long-term forgetting (ALF), is characterized by an increased rate of memory loss over extended intervals despite intact initial learning and recall (Mameniskiene et al., 2020). As standard memory tests typically assess recall only after short delays, ALF may remain undetected in clinical practice. This phenomenon has been well-documented in temporal lobe epilepsy (Elliott et al., 2014). More recently, it has also been identified in other neurological conditions such as traumatic brain injury (TBI), transient ischemic attack (TIA) and minor stroke as well as limbic encephalitis (Butler et al., 2019). Weston et al. (2018) were also able to detect ALF in presymptomatic carriers of Alzheimer’s disease mutations by analyzing the ratio of 7-day to 30-minute delayed recall performance on list, story, and figure memory tests. The same method was applied by Stalter et al. (2024) in a small cohort of mildly impaired MS patients, where ALF was also demonstrated, particularly affecting verbal memory. However, the presence and clinical relevance of ALF in larger, more heterogeneous MS populations remain unclear.

Additionally, MS patients frequently report subjective memory impairment, which often lacks objective verification (Krch et al., 2011). Dementia research has shown that cognitive decline occurs on a continuum, ranging from subclinical changes, to subjective cognitive deficits, then to mild cognitive impairment, and ultimately to clinical evident dementia (An et al., 2024). Stalter et al. (2024) suggested that ALF may provide an objective correlate for self-reported memory difficulties and fatigue.

This study aims to (I) determine the presence of ALF in a larger, clinically diverse cohort of MS patients, (II) explore the relationship between ALF and patient-reported outcomes, including SMI, fatigue, depression, and sleep disturbances, and (III) evaluate the diagnostic performance of the newly established ALF assessment method, using the RAVLT and the WMS-IV Logical Memory subtest, to discriminate subjective memory impairment.

## 2 Materials and methods

### 2.1 Selection and description of participants

Patients with MS were recruited at the Evangelical Hospital and its university outpatient clinic in Oldenburg, Germany, between 2020 and 2025. Eligible participants were ≥ 18 years, native German speakers, and diagnosed with MS (RRMS, SPMS, or PPMS) according to the 2017 revised McDonald criteria (Thompson et al., 2018). Exclusion criteria comprised the use of any CNS-active medication, including corticosteroids, within six weeks prior to enrollment, as well as any neurological or psychiatric comorbidity besides MS. In addition, a Montreal Cognitive Assessment (MoCA) score < 26 and a Beck Depression Inventory II (BDI - II) score ≥ 20 were exclusion criteria (Herzberg et al., 2008; Nasreddine et al., 2005). Participants who did not complete the assessment were also excluded from the analyses. Healthy controls were matched for age, sex, and years of education and were, along with the given criteria, required to have no neurological and psychiatric conditions. Demographical information was obtained via self-report. Written informed consent was obtained from all participants prior to enrollment. The study was approved by the Ethics Committee of the Carl von Ossietzky University in Oldenburg, Germany (approval number 2024-163) and is registered in the German Clinical Trials Register (DRKS00035204).

### 2.2 Data collection and measurements

Verbal memory was assessed using Rey’s Auditory Verbal Learning Test (RAVLT) and the Logical Memory subtest of the Wechsler Memory Scale-IV (WMS-IV).

The RAVLT contains a list of 15 words that the participant learns within 5 trials. After an interference list, the memorized terms are reproduced (0 – 15 points) (Bean, 2018). The WMS-IV Logical Memory subtest consists of two stories from which as many details as possible should be remembered (0 – 50 points) (Lepach and Petermann, 2012). A delayed recall follows after 30 minutes. To assess ALF, an additional 7-day-delayed recall was obtained via telephone interview. To avoid influencing the 7-day-delayed recall performance, the content of the interview was not disclosed. Primary outcomes were the 7-day/ 30-minute delayed recall ratios for the RAVLT (*Q*_RAVLT_) and WMS-IV Logical Memory (*Q*_WMS_), following the methodology used in earlier studies (Stalter et al., 2024; Weston et al., 2018). Only complete cases were included in the analyses.

Additional neurocognitive and neuropsychiatric measures included the MoCA, Symbol Digit Modalities Test (SDMT), and questionnaires including the Fatigue Impact Scale (FIS), BDI - II, Pittsburgh Sleep Quality Index (PSQI) and Epworth Sleepiness Scale (ESS) (Buysse et al., 1989; Häuser et al., 2003; Herzberg et al., 2008; Johns, 1991; Nasreddine et al., 2005; Smith, 1973). Subjective memory difficulties were evaluated with the Everyday Memory Questionnaire (EMQ) and two German questionnaires focusing on memory and attention deficits in everyday life (FEAG, FEDA) (Hupfeld, 2009; Sunderland et al., 1983; Zimmermann et al., 1991). In accordance with previous work, self-reported memory assessment was completed using two different methods. The participants were asked to report whether they currently experienced increased forgetfulness (yes/no) and to rate their perceived forgetfulness on a scale from 0 (not forgetful) to 100 (very forgetful) (Stalter et al., 2024).

### 2.3 Statistical methods

Differences in MS patients and healthy controls were analyzed using Chi-square or Fisher’s exact test in categorial demographic variables. Continuous demographic variables, neuropsychological, and questionnaire data were compared using the Mann-Whitney-U-test. For the two primary outcomes, *Q*_RAVLT_ and *Q*_WMS_, Bonferroni correction was applied (α = 0.025). Linear regression models were constructed using Q_RAVLT_ or Q_WMS_ as dependent variables; each questionnaire measure (FIS, BDI-II, PSQI, ESS, EMQ, FEAG, FEDA), Expanded Disability Status Scale (EDSS), disease duration (time since diagnosis), and age were entered as independent variables in separate models. Regression analyses were considered exploratory. The combined ALF score was calculated as the mean of the Q_RAVLT_ and the Q_WMS_ score. Receiver operating characteristic (ROC) analysis of the combined ALF score was conducted to evaluate diagnostic accuracy in discriminating subjective memory impairment, assessed by asking patients whether they had recently perceived themselves as more forgetful, reporting area under the curve (AUC), 95% confidence interval, optimal cut-off (based on the Youden Index), sensitivity, and specificity. All analyses and graphical visualizations were performed in RStudio (Version 2024.04.2+764) (Posit, 2024).

## 3 Results

### 3.1 Sample characteristics

Seventy-six MS patients and 67 healthy controls were initially enrolled in the study. After exclusion of participants, due to failure to complete the delayed recall assessment (n=3), MoCA-Score < 26 (n=8), BDI-II-score ≥ 20 (n=4), or treatment with CNS-active medication (n=2), 62 MS patients and 65 healthy controls (HC) were included for the analyses. Demographic and clinical characteristics of the study population are summarized in Table 1. Additional information on the disease-modifying medication used by MS patients can be found in Table A.1. Demographic characteristics did not differ significantly between both groups. Age, sex distribution, and MS phenotypes in the patient sample were roughly consistent with epidemiologic characteristics reported for MS populations (Multiple Sclerosis International Federation, 2020).

**Table 1.**
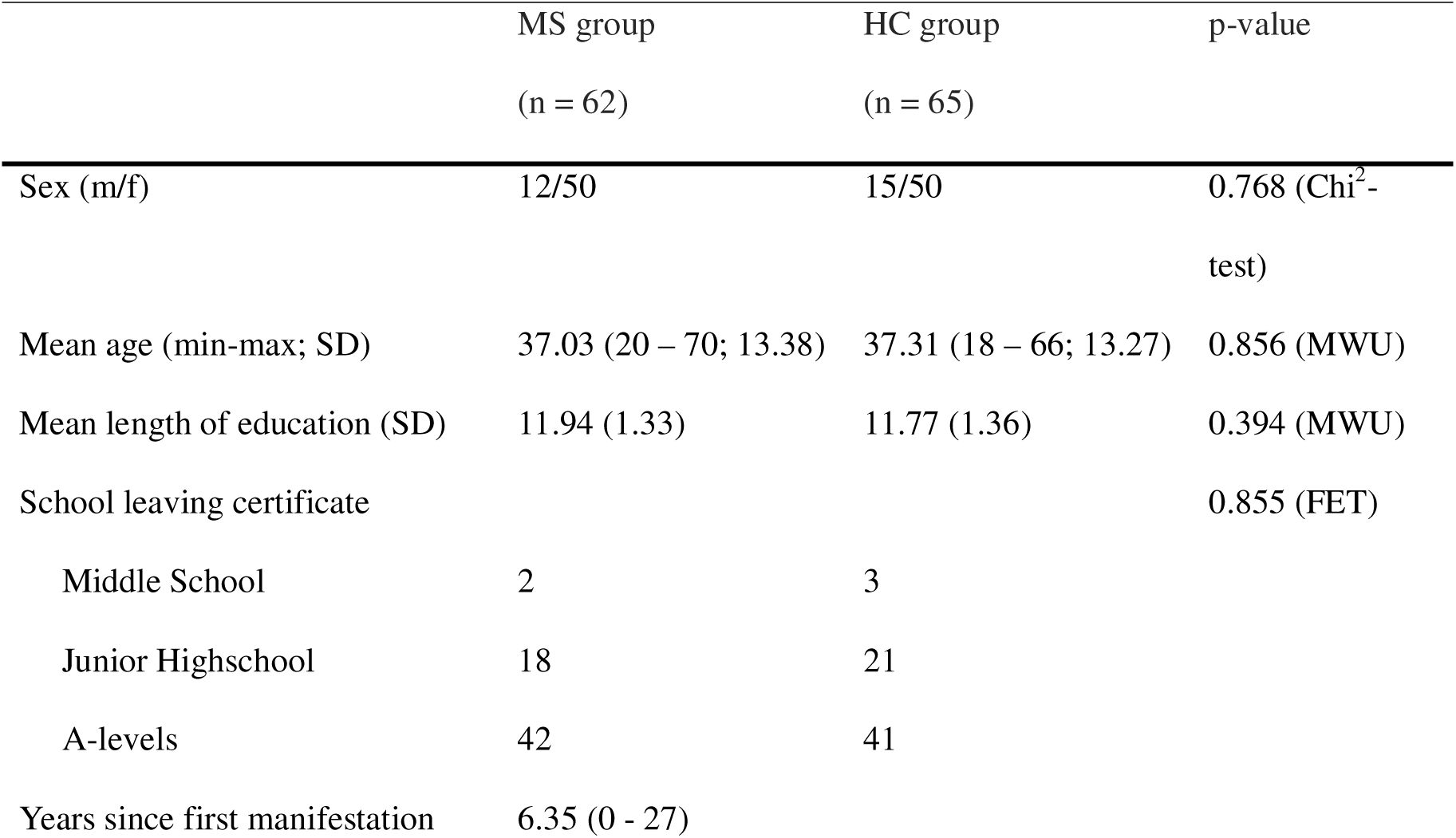
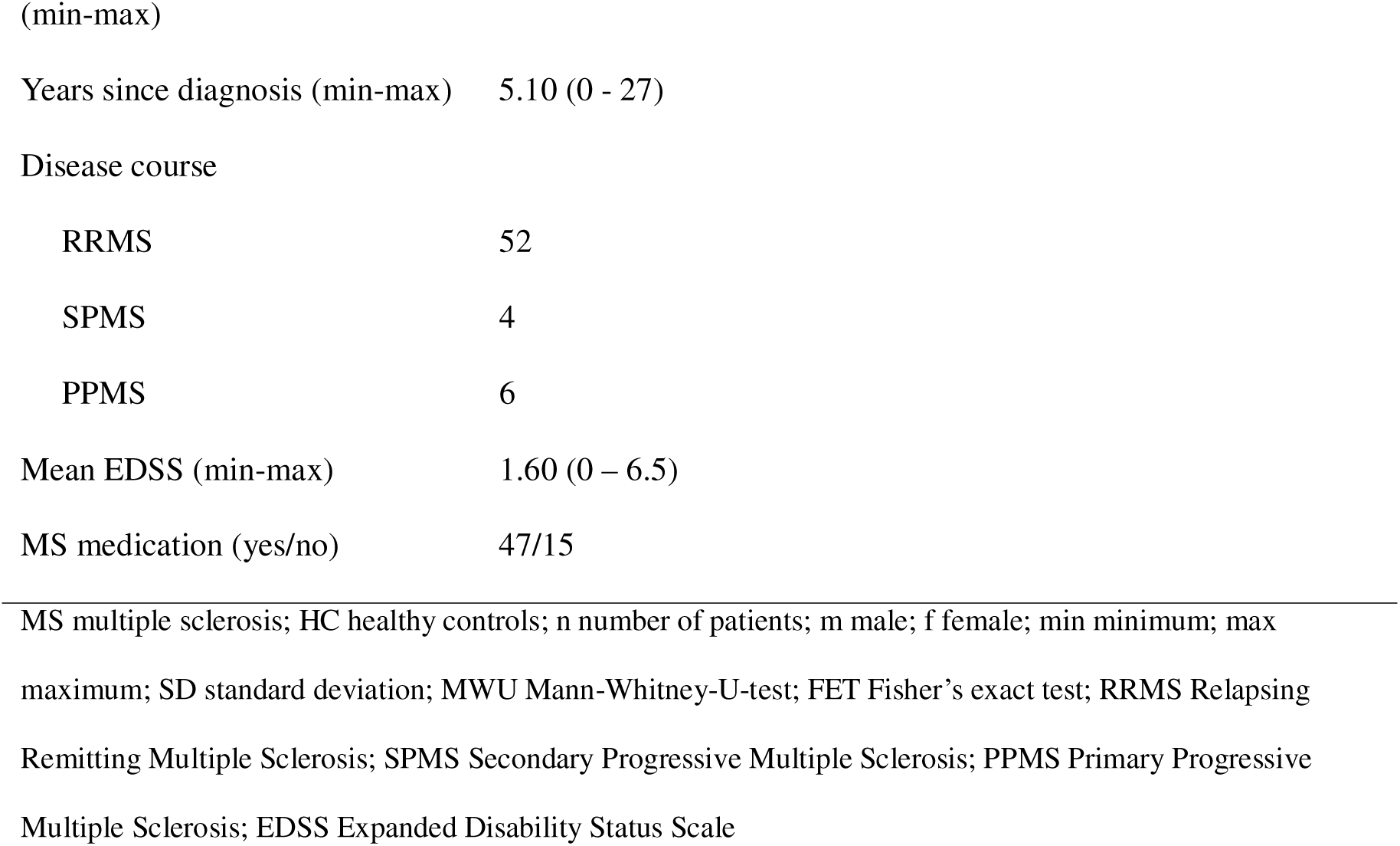
Demographical and clinical characteristics.

### 3.2 Primary outcomes

Across both verbal memory tests, MS patients demonstrated significantly greater ALF than controls as measured with the described quotient. MS patients recalled fewer words after 7 days, resulting in a lower Q_RAVLT_ compared with HC (0.64 [95% CI 0.59–0.69] vs. 0.78 [0.73–0.82], p < 0.001). A similar pattern was observed for story recall: MS patients reproduced fewer story elements after 7 days, yielding lower Q_WMS_ scores (0.79 [95% CI 0.74–0.84] vs. 0.95 [0.90–1.0], p < 0.001). Immediate recall and 30-minute delayed recall did not differ between groups for either test. Complete results for the RAVLT and WMS-IV Logical Memory subtest are presented in Table 2, and performance trajectories across delay intervals are visualized in Figure 1. Initial Trial-by-trial learning curves for the RAVLT did not differ between groups, as shown in Figure 2. Mean scores across RAVLT trials are provided in Table A.2.

**Figure 1.**
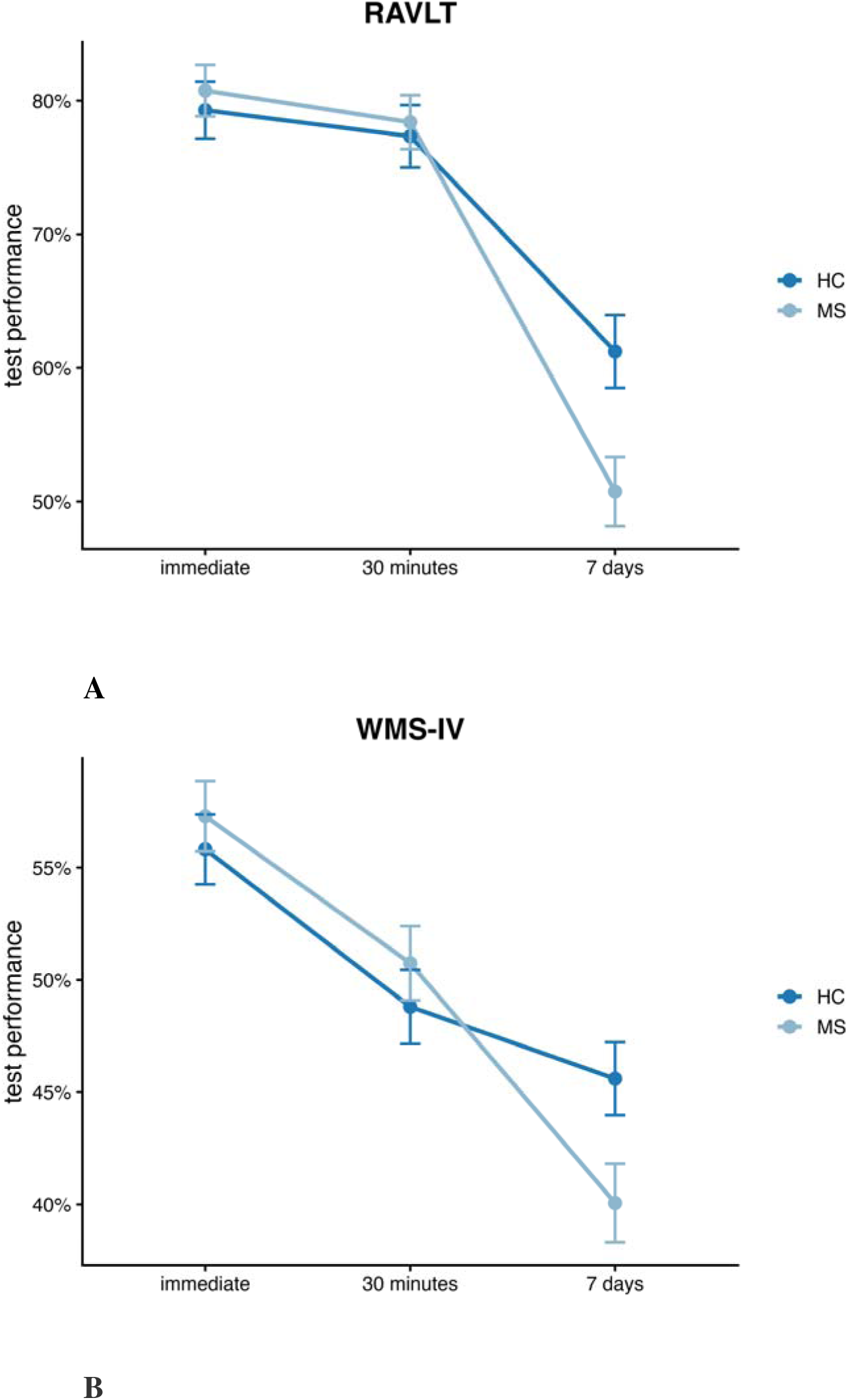
Performance trajectories across delay intervals. Mean (± SE) test performance (%) for the RAVLT (A) and WMS-IV Logical Memory subtest (B) in each group at the various test times. SE standard error; RAVLT Rey’s Auditory Verbal Learning Test; WMS Wechsler Memory Scale; HC healthy controls; MS multiple sclerosis

**Figure 2.**
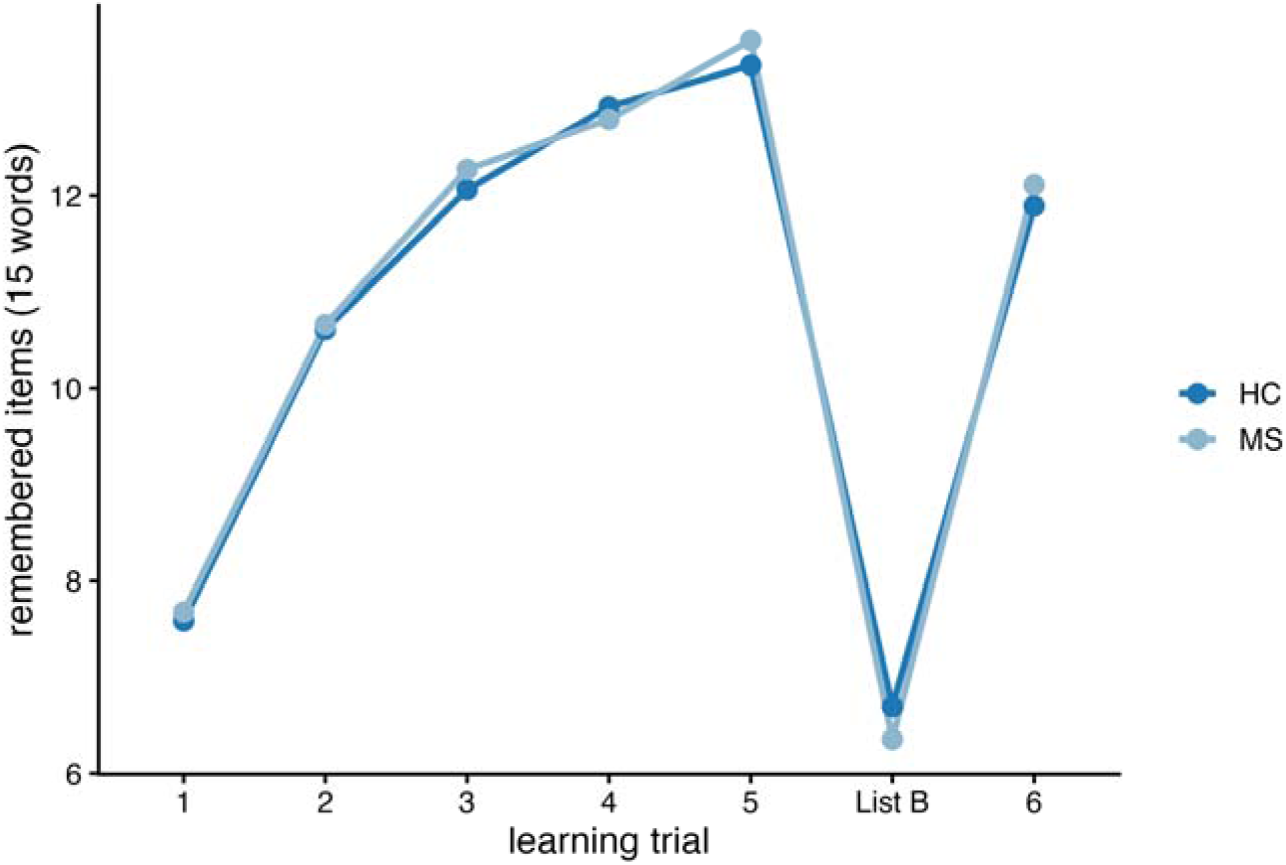
Trial-by-trial learning curves of the RAVLT. Learning curves of the mean number of remembered items in each group for the RAVLT. RAVLT Rey’s Auditory Verbal Learning Test; HC healthy controls; MS multiple sclerosis

**Table 2.** ALF test results.

|  | MS group<br>(n = 62) | HC group<br>(n = 65) | p-value |
| --- | --- | --- | --- |
| **Results after initial testing** |  |  |  |
| RAVLT initial Score<br>(%, min-max) | 80.75 (40 – 100) | 79.28 (26.67 - 100) | 0.924 (MWU) |
| WMS-IV initial Score<br>(%, min-max) | 57.29 (28 - 84) | 55.82 (28 - 84) | 0.457 (MWU) |
| **Results after 30 minutes** |  |  |  |
| RAVLT 30-minute-score<br>(%, min-max) | 78.39 (46.67 - 100) | 77.33 (20 - 100) | 0.923 (MWU) |
| WMS-IV 30-minute-score<br>(%, min-max) | 50.74 (18 - 80) | 48.80 (18 - 78) | 0.508 (MWU) |
| **Results after 7 days** |  |  |  |
| RAVLT 7-day-score<br>(%, min-max) | 50.75 (6.67 – 93.33) | 61.23 (13.33 – 100) | 0.004 (MWU) |
| WMS-IV 7-day-score<br>(%, min-max) | 40.06 (10 – 78) | 45.60 (12 – 70) | 0.007 (MWU) |
| $Q_{\text{RAVLT}}$ (min-max) | 0.64 (0.13 – 1.13) | 0.78 (0.29 – 1.22) | < 0.001 (MWU) |
| $Q_{\text{WMS}}$ (min-max) | 0.79 (0.19 – 1.12) | 0.95 (0.32 – 1.50) | < 0.001 (MWU) |

MS multiple sclerosis; HC healthy controls; n number of patients; RAVLT Rey's Auditory Verbal Learning Test;
WMS Wechsler Memory Scale; min minimum; max maximum; MWU Mann-Whitney-U-test

### 3.3 Additional neurocognitive test results and self-reported symptoms

Global cognition (MoCA) and information processing speed (SDMT) did not show significant differences between both groups. MS patients self-reported significantly greater fatigue (FIS) and poorer sleep quality (PSQI) than control participants. Subjective memory complaints, as assessed using the EMQ, FEAG, FEDA, and the two self-ratings of forgetfulness (binary and 0-100 scale), were also significantly elevated in MS patients, with exception of the FEDA. All results for additional neurocognitive tests and questionnaire scores are summarized in Table 3.

**Table 3.**
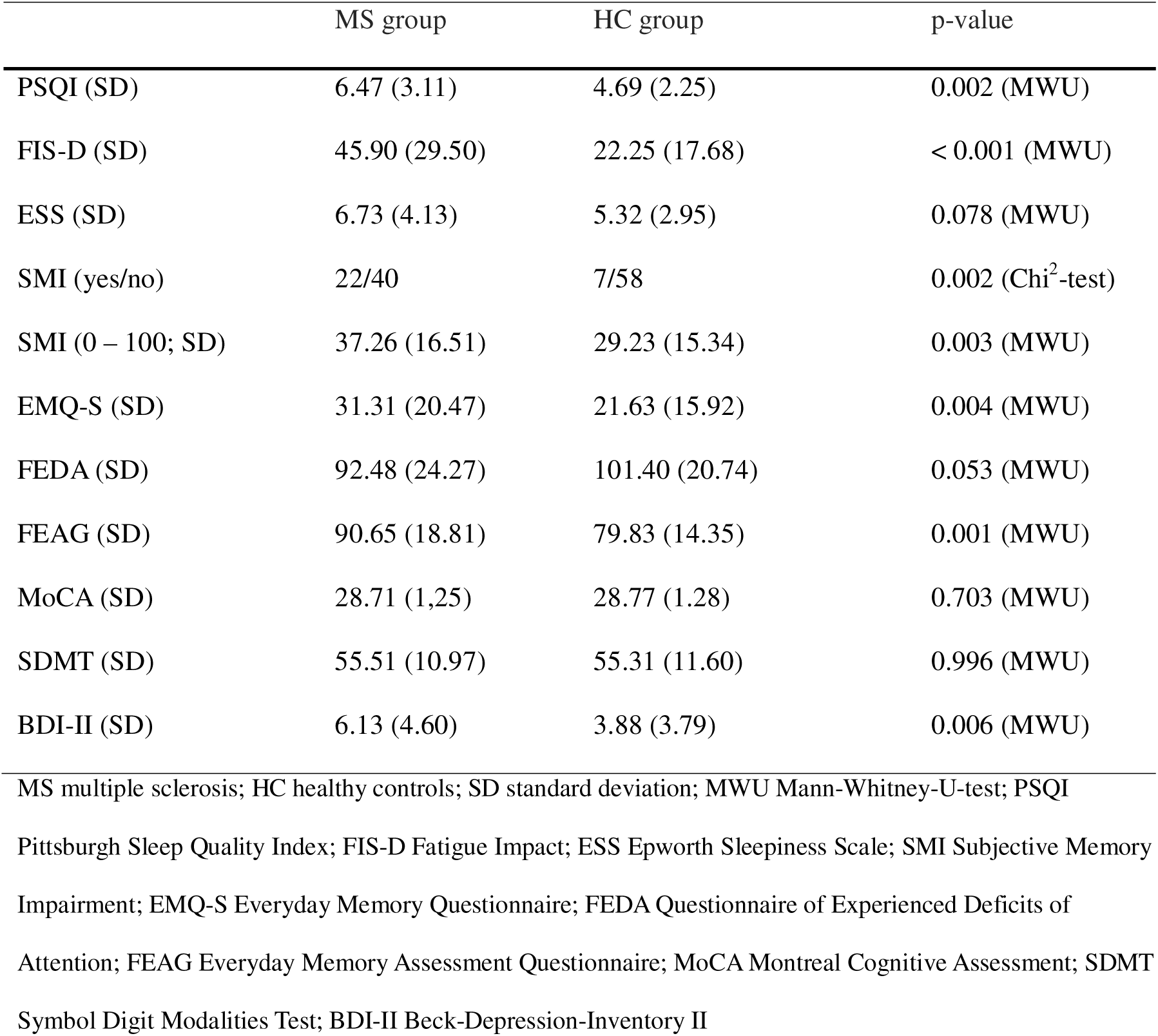
Questionnaires and other test results.

### 3.4 Regression analyses

Within the MS group, fatigue and age emerged as possible predictor of Q_RAVLT_ performance. For Q_WMS_, significant predictors included fatigue, questionnaires assessing everyday memory and attentional difficulties (FEAG, FEDA, EMQ), greater disability (EDSS), longer disease duration, and age. No significant associations were identified within the HC group. Detailed regression coefficients and model statistics are provided in Table 4.

**Table 4.** Linear regression models in the multiple sclerosis group.

| Outcome | Predictor | $\beta$ | p-value | 95% CI | $R^2$ | $R^2$<br>adjusted |
| --- | --- | --- | --- | --- | --- | --- |
| $Q_{RAVLT}$ | FIS-D | -0.002 | 0.006 | -0.004 to -0.001 | 0.117 | 0.102 |
| $Q_{WMS}$ | FIS-D | -0.004 | < 0.001 | -0.005 to -0.002 | 0.288 | 0.276 |
| $Q_{WMS}$ | EMQ | -0.003 | 0.007 | -0.006 to -0.0009 | 0.116 | 0.101 |
| $Q_{WMS}$ | FEAG | -0.004 | < 0.001 | -0.007 to -0.002 | 0.177 | 0.164 |
| $Q_{WMS}$ | FEDA | 0.005 | < 0.001 | 0.003 to 0.006 | 0.319 | 0.308 |
| $Q_{WMS}$ | EDSS | -0.068 | 0.001 | -0.106 to -0.030 | 0.181 | 0.166 |
| $Q_{WMS}$ | Time since diagnosis | -0.010 | 0.021 | -0.018 to -0.002 | 0.086 | 0.070 |
| $Q_{RAVLT}$ | Age | -0.005 | 0.006 | -0.009 to -0.001 | 0.118 | 0.104 |
| $Q_{WMS}$ | Age | -0.008 | < 0.001 | -0.011 to -0.004 | 0.273 | 0.261 |

RAVLT Rey's Auditory Verbal Learning Test; WMS Wechsler Memory Scale; FIS-D Fatigue Impact Scale;
EMQ Everyday Memory Questionnaire; FEAG Everyday Memory Assessment Questionnaire; FEDA
Questionnaire of Experienced Deficits of Attention; EDSS Expanded Disability Status Scale; CI confidence interval

### 3.5 Discriminatory accuracy

ROC analysis demonstrated moderate discriminatory accuracy of the combined ALF score for detecting subjective memory impairment (AUC = 0.735, 95% CI 0.585 - 0.884). The optimal cut-off value, determined using the Youden Index, was 0.71, yielding a sensitivity of 0.727 and a specificity of 0.725. The corresponding ROC curve is shown in Figure 3.

**Figure 3.**
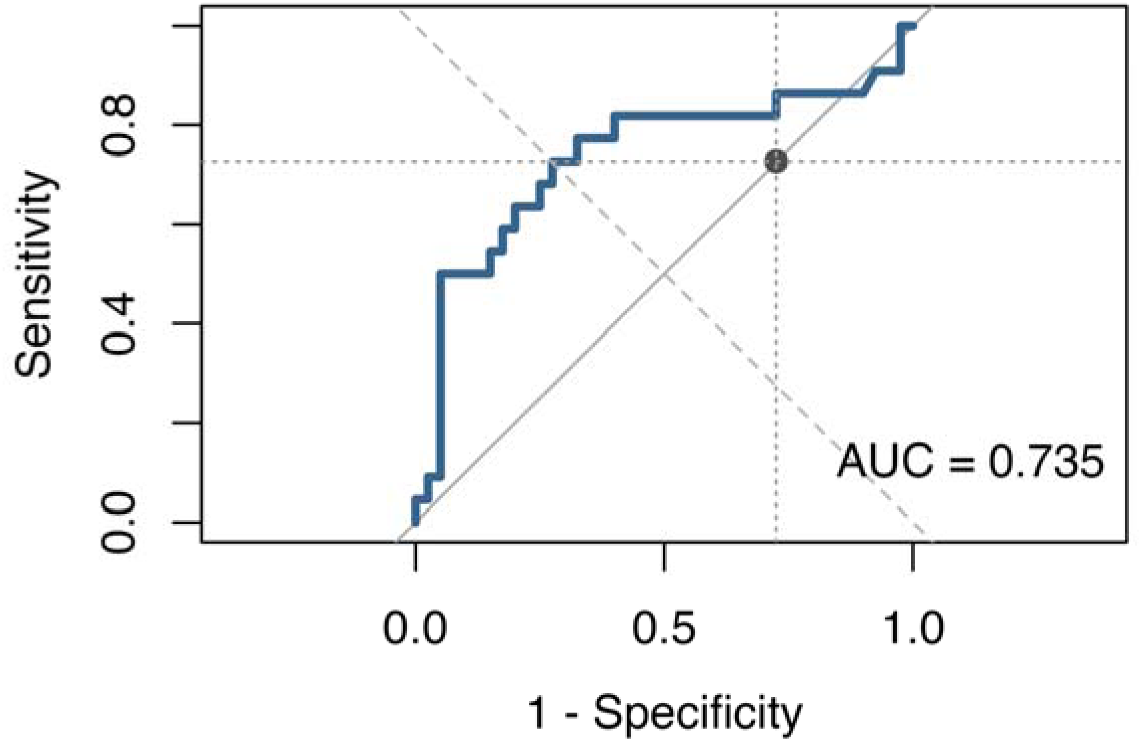
ROC curve of the combined ALF score for detecting subjective memory impairment. Receiver operating characteristic (ROC) curve for the combined ALF score, illustrating the diagnostic accuracy in discriminating between presence vs. absence of subjective memory impairment. Sensitivity is plotted against the false-positive rate (1-specificity). The diagonal dotted line represents chance performance. The dot indicates the optimal cut-off based on the Youden Index. AUC area under the curve

## 4 Discussion

In this study, we administered the RAVLT and the WMS-IV Logical Memory subtest to a heterogeneous group of MS patients and matched healthy controls to replicate previously reported evidence of ALF in MS in a larger group of patients. In addition, we used linear regression models to identify potential predictors of ALF within the patient group, and we examined the diagnostic performance of the RAVLT and the WMS-IV Logical Memory subtest for discriminating subjective memory impairment.

Consistent with the findings of Stalter et al. (2024), we observed significantly lower ALF scores in MS patients when assessed with the word list (Q_RAVLT_). Beyond replicating these results, we were able to demonstrate significantly reduced ALF scores in the story-learning test (Q_WMS_). For both neuropsychological tasks, there was no difference between patients and controls in terms of immediate and short-delay recall or learning ability. These findings indicate that ALF in MS is caused by a problem in late consolidation rather than during initial encoding or early consolidation.

By applying these tests to a broader and clinically more representative MS sample, we were able to increase the generalizability of previous findings and thereby confirm our first hypothesis. Our study shows that ALF affecting verbal memory is present in a patient population that resembles real-world clinical demographics, with the predominance of female patients and a relapsing-remitting disease course (Multiple Sclerosis International Federation, 2020). The materials used for this study met the requirements that Baddeley et al. (2019) set for ALF assessment materials. Both tests provide reliable results when repeated, without requiring long learning periods. As the participants were not informed about the delayed recall assessment, strategic differences were minimized, and the tests also allowed for remote retesting. Furthermore, we mostly followed the recommendations proposed by Elliott et al. (2014) to ensure the quality of ALF assessment. Patients and controls were matched for age and intellectual ability, recall and recognition were tested, and distraction tasks ensured that the information was transferred to long-term memory. Ceiling and floor effects were reduced by matching initial learning, and practice of the test material prior to delayed retrieval was prevented.

With this study we extended the findings on ALF, suggesting that this specific cognitive impairment is also relevant in MS, aligning with findings from other neurological conditions (Butler et al., 2019). ALF has previously been linked to disruptions in hippocampal-neocortical interaction and long-term consolidation mechanisms in patients with temporal lobe epilepsy (TLE) (Audrain and McAndrews, 2019; Mameniskiene et al., 2020). While forgetting over short intervals is associated with hippocampal changes, its contribution to ALF remains poorly understood (Audrain and McAndrews, 2019). It is well established that the hippocampus is primarily involved in the early phase of memory formation, while neocortical structures become increasingly relevant over time (Mameniskiene et al., 2020). Audrain and McAndrews (2019) demonstrated that ALF in TLE patients is associated with reduced resting-state connectivity between the anterior hippocampus and the lateral temporal cortex. This finding supports the assumption that ALF cannot be completely explained by hippocampal changes; rather, it reflects impaired interactions within hippocampal-neocortical networks.

Disconnection of the hippocampus from different brain networks, as well as focal hippocampal damage, is also known to be a contributing factor to cognitive impairment in MS (Rocca et al., 2018). These parallels suggest that ALF in MS may arise from similar disruptions in long-term consolidation pathways as those described in other neurological conditions. Inflammatory demyelination and neurodegenerative processes may interfere with the transfer of acquired information into neocortical storage sites. Future studies that extend the current methods by integrating structural and functional imaging could help clarify the neurobiological mechanisms underlying ALF in MS and determine whether ALF may be useful as an early cognitive marker for subtle network dysfunction.

When considering potential factors associated with ALF, previous research has identified fatigue and self-perceived forgetfulness (rated on a 0-100 scale) as relevant variables (Stalter et al., 2024). Using regression analyses, we were able to replicate the findings on fatigue in our study, indicating that patients with greater fatigue show lower ALF scores. Kinsinger et al. (2010) demonstrated that fatigue influences subjective, but not objective measures of neuropsychological functioning. Combined with our findings, this may suggest that fatigue affects long-term memory formation and consolidation rather than short-term memory, which is typically assessed in standard neuropsychological testing. The comorbidity of fatigue and ALF could be explained because both are dependent on sleep quality, which is often impaired in MS patients (Carnicka et al., 2015).

In addition, we observed that greater disability, as measured by the EDSS, longer disease duration, and age were associated with lower ALF scores. Prior evidence has shown that longer disease duration and older age relate to cognitive impairment in multiple sclerosis, whereas the association with EDSS was only a trend (Wu et al., 2025). Given that age is considered to be an important determinant of ALF, age-related effects may not be disease-specific (Elliott et al., 2014). Overall, these findings suggest that the cumulative effects of neurodegeneration may also contribute to impaired long-term memory consolidation.

Regarding subjective memory impairment, our results also point to an association with ALF. In our study, this relationship was identified using self-report questionnaires (FEAG, FEDA, EMQ). Stalter et al. (2024) reported comparable findings, although they found associations with memory complaints using a 0-100 self-rating scale. Our findings suggest that the measures used to assess perceived memory difficulties may need to be further improved to accurately predict objective memory impairment.

In summary, ALF testing may provide an objective complement to subjective memory complaints and fatigue-related difficulties. However, the questionnaires, rating scales, and binary questions employed in the present study appear insufficient for a comprehensive assessment of subjective memory impairment.

Regarding the diagnostic performance in discriminating subjective memory impairment, ROC analysis indicated moderate accuracy for the combined ALF score. Thus, the long-delay recall extensions of the RAVLT and the WMS-IV Logical Memory subtest appear suitable for identifying memory impairments that remain undetected in short-delay recall assessments. Given their sufficient sensitivity for detecting ALF, these methods allow the identification of individuals who may benefit from more extensive neuropsychological testing.

Acknowledging the fact that cognitive decline progresses continuously, with subjective memory complaints preceding mild cognitive impairment and eventually dementia, ALF measures may offer an approach for earlier detection and enable interventions that may positively influence symptom progression. This finding highlights the potential clinical value of detecting subtle memory deficits that standard neuropsychological tests may not recognize. As we are using a cross-sectional design, we cannot provide information on the evolution of ALF over time. Additional longitudinal studies could help to determine whether ALF can also predict disease progression and future cognitive decline in MS patients.

Some limitations should be considered when interpreting our findings. We assessed ALF exclusively in the domain of verbal memory. Elliott et al. (2014) recommend that the evaluation should include both verbal and nonverbal memory modalities. Although visuospatial memory ALF has been observed across different patient populations, it has not yet been demonstrated in patients with MS (DeLuca et al., 1998; Mameniskiene et al., 2020; Rao et al., 1991; Weston et al., 2018). One possible explanation is the negative association of the visuospatial memory with the EDSS as found by Artemiadis et al. (2020), since the EDSS in our study was rather low. In their study, Stalter et al. (2024) used Rey’s complex figure test to assess ALF in visuospatial memory. However, results did not differ between MS patients and controls, as both groups performed poorly on the 7-day delayed recall (Stalter et al., 2024). Given these findings, future studies should consider using alternative methods to investigate ALF in visuospatial memory.

Furthermore, the methods used to detect subjective memory impairment may limit the interpretation of our findings, as there is no standardized method for assessing self-experienced forgetfulness (Abdulrab and Heun, 2008). Abdulrab and Heun (2008) recommend the specification of time-related details, providing examples, and obtaining confirmation through a relative. Future studies therefore should consider alternative measures for subjective cognitive deterioration, as current questionnaires and visual analogue scales may not be sufficient.

Other limitations involve the use of medication in the patient group. While we defined the use of medication with a proven effect on CNS-function as an exclusion criterion, 47 of the 62 patients used nonsteroid immune suppressive MS-medication. The effect of these drugs on memory function has not yet been sufficiently investigated.

MS is the most common cause of non-traumatic disability in young adults (Tullman, 2013). Cognitive deficits, such as objectively detectable and subjectively perceived memory impairments, contribute substantially to impairments in professional and private life (Amato et al., 2006). Unlike many other chronic neurological diseases, MS usually manifests itself in young, active people, meaning that the symptoms of the disease accompany those affected for a long time (Strober et al., 2012). Paving the way for reducing these relevant effects on patients is therefore the primary goal of this study. This work aims to contribute to improved neuropsychological evaluation by testing a methodology for the reliable ALF assessment, which has already been successful in previous studies (Stalter et al., 2024; Weston et al., 2018). Establishing sensitive methods for the detection of ALF may provide access to therapeutic interventions for memory impairment at an early stage, thereby positively influencing the course of the disease.

## Ethics approval

This study was approved by the Ethics Committee of the Carl von Ossietzky University in Oldenburg, Germany (approval number 2024-163) and was conducted in accordance with the principles of the Declaration of Helsinki.

## Data availability

Data supporting this study’s findings are available from the corresponding author upon reasonable request.

## Author contributions

CJ: conceptualization, methodology, validation, formal analysis and investigation, resources, data curation, writing - original draft preparation, writing - review and editing, visualization, project administration, funding acquisition. JS: conceptualization, methodology, validation, investigation, data curation, formal analysis and investigation, resources, data curation, writing - original draft preparation, writing - review and editing, visualization, funding acquisition. SR: validation, investigation, resources, writing - review and editing, project administration, funding acquisition. KW: conceptualization, methodology, validation, formal analysis, investigation, resources, data curation, writing - original draft preparation, writing - review and editing, visualization, supervision, project administration, funding acquisition

## Acknowledgements

We would like to thank the Hertie Foundation for funding this study.

