## Supplementary material for "Accelerated long-term forgetting as an objective marker of subjective memory impairment in multiple sclerosis": Table A.

**Table A.1** Disease - modifying medication in multiple sclerosis patients

| Medication name | MS patients (n, %) |
| --- | --- |
| Ocrelizumab | 14 (22.58) |
| Ofatumumab | 10 (16.13) |
| Dimethyl fumarate | 9 (14.52) |
| Glatiramer acetate | 7 (11.29) |
| Teriflunomide | 3 (4.84) |
| Natalizumab | 2 (3.23) |
| Interferon-beta 1a | 1 (1.61) |
| Diroximel fumarate | 1 (1.61) |

MS multiple sclerosis; n number of patients

**Table A.2** Mean scores across RAVLT trials

|  | MS group  (mean, SD) | HC group  (mean, SD) | p-value  (MWU) |
| --- | --- | --- | --- |
| Trial 1 | 7.68 (1.89) | 7.58 (1.80) | 0.853 |
| Trial 2 | 10.66 (2.16) | 10.61 (2.16) | 0.773 |
| Trial 3 | 12.27 (1.88) | 12.06 (1.98) | 0.530 |
| Trial 4 | 12.79 (1.80) | 12.92 (1.84) | 0.646 |
| Trial 5 | 13.61 (1.51) | 13.35 (1.72) | 0.407 |
| Interference List B | 6.35 (1.98) | 6.69 (2.06) | 0.335 |
| Trial 6 (Post - Interference Recall) | 12.11 (2.27) | 11.89 (2.57) | 0.924 |

RAVLT Rey’s Auditory Verbal Learning Test; MS multiple sclerosis; HC healthy controls; SD standard deviation; MWU Mann-Whitney-U-test
